# Difficult-to-treat rheumatoid arthritis (D2T RA): clinical issues at early stages of disease

**DOI:** 10.1101/2022.11.04.22281931

**Authors:** Leticia Leon, Alfredo Madrid-García, Patricia Lopez-Viejo, Isidoro González-Álvaro, Marta Novella-Navarro, Dalifer Freites Nuñez, Zulema Rosales, Benjamin Fernandez-Gutierrez, Lydia Abasolo

**Affiliations:** Instituto de Investigacion Sanitaria San Carlos (IdISSC). Rheumatology Department. Hospital Clínico San Carlos. Madrid, Spain; Health Sciences, Universidad Camilo Jose Cela. Madrid,Spain; Rheumatology Department. Hospital Clínico San Carlos. Madrid, Spain; Rheumatology Department. Hospital Universitario La Princesa. Madrid, Spain 5. Rheumatology Department. Hospital Universitario La Paz. Madrid, Spain

## Abstract

**Objectives:** Most studies on difficult-to-treat rheumatoid arthritis (D2T RA) have focused on established RA. Here, we analyze whether disease activity in the early stages of RA could influence progression to a D2T RA under real-life conditions. Other clinical and treatment-related factors were also analyzed.

**Methods:** A longitudinal multicenter study of RA patients was conducted from 2009 to 2018. Patients were followed up until January 2021. D2T RA was defined based on EULAR criteria (treatment failure, signs suggestive of currently active/progressive disease, and management being perceived as problematic by the rheumatologist and/or patient). The main outcome was disease activity in the early stages. The covariates were sociodemographic, clinical, and treatment-related factors. We ran a multivariable logistic regression analysis to investigate risk factors associated with progression to D2T RA. Weighting techniques were also applied to balance data.

**Results:** The study population comprised 631 patients and 35 developed D2T RA. At the time of diagnosis, the D2T RA group were younger, with a higher disability, DAS28 score, tender joint count and pain scores. In our final model, DAS28 was not statistically significantly associated with D2T RA. No differences were found between groups for therapy. Disability was independently associated with D2T RA (OR: 1.50; p=0.02).

**Conclusions:** In this cohort of patients newly diagnosed with RA, our results do not allow us to prove the influence of active disease according to DAS28. However, we did find that patients with elevated initial disability scores are more likely to develop D2T RA regardless of other factors.

**What is already known on this topic:** Despite T2T and the availability of a range of advanced therapies, D2T RA remains a relevant clinical problem. Evidence for the D2T RA population have focused on established RA. The aim of our study was to analyze whether disease activity at diagnosis could influence progression to D2T RA under real-life conditions.

**What this study adds:** We did find that patients with elevated initial disability scores are more likely to develop D2T RA regardless of other factors.

**How this study might affect research, practice or policy:** The implementation of more effective strategies in the early stages of the disease and focused on the most influential factors, including severe disability, may change disease course and prevent D2T RA.

## INTRODUCTION

The main therapeutic objective in rheumatoid arthritis (RA) is to prevent joint damage and disability. Treatment seeks to achieve and maintain “remission” or, at least, minimal disease activity. Recent changes in the management of RA have improved the clinical course of the disease (1). The “window of opportunity” concept (2)(3)(4) underscores the need to initiate aggressive treatment early in the disease process and thus improve prognosis. Intensification of treatment, dose optimization, and the “treat to target” strategy (5)(6)(7)(8) are also key goals in the management of affected patients. Remarkable progress in treatment has been made with both biological therapies and JAK-STAT pathway inhibitors.

Recommendations on the management of RA have come to include continuous supervision and monitoring of disease course and associated comorbidity (9)(10), thus enabling remission and low disease activity in clinical practice (1). The prevalence of remission in RA patients is reported to be around 30% (11)(12). Prognosis has improved in recent decades and may reflect early diagnosis and more accurate treatment rather than a change in the characteristics of the process itself (13).

Despite these advances, there are patients with poor disease evolution, reflecting the fact that RA is a heterogeneous disease in terms of severity, clinical course, and response to treatment, requiring a complex management. A European League Against Rheumatism (EULAR) Task Force recently defined the concept of “difficult-to-treat” (D2T) RA as persistent symptoms and/or signs after failure of at least two biological or targeted synthetic disease modifying anti-rheumatic drugs (b/tsDMARDs), each of which has a different mechanism of action(14). D2T RA encompasses not only uncontrolled inflammatory disease, but also wider contextual factors such as chronic pain and fatigue, as well as comorbidities, recurrent infections, and treatment-limiting adverse events. Up to 20% of RA patients develop difficult-to-treat disease (15)(16)(17).

Although the mechanisms leading to D2T RA are varied and not fully understood, they can be classified into two types, namely, multi-drug resistance and difficulties in intensifying treatment(15)(18). Multi-drug resistance can be ascribed to the underlying immune disorders of RA and environmental factors such as smoking, pharmacogenetics, and drug immunogenicity. Difficulties in intensifying treatment include comorbidities, poor adherence to medication, financial status, or the patient’s reluctance to intensify treatment.

In a recent study of more than 1,700 patients, 10% of all RA patients did not achieve remission or low disease activity despite intensive treatment in real clinical practice (19). The prevalence of DT2 RA is assumed to range between 3% and 17% according to the series reviewed(15)(20). The main characteristics of patients with D2T RA are younger age, high 28-joint Disease Activity Score (DAS28), comorbidity with fibromyalgia, marked disability according to the Health Assessment Questionnaire (HAQ), high visual analog scale (VAS) score, poorer quality of life, increased number of DMARDs, higher doses of glucocorticoids, patient desire to intensify treatment, and greater consumption of healthcare resources(18). Evidence for the EULAR-defined D2T RA population is limited and based mainly on definitions that do not take into account features of active disease and the perception of challenging RA by the clinician and/or patient (21), focusing mainly on the first of them (failure of at least two b/tsDMARDs). In addition, almost all patients included have long-standing RA. A recent study found that prognosis was difficult in patients with D2T RA who receive MTX ≤⍰8.6 mg/week or who receive glucocorticoids. Therefore, focusing on the most effective treatments from the onset of the disease may prevent increases in its frequency (22).

Knowledge about why some patients develop D2T RA could be improved. The few studies performed are cross-sectional, regardless of the duration of RA. Consequently, to identify patients at risk, it would be interesting to identify modifiable associated factors at early stages. The aim of our study was to analyze whether disease activity at diagnosis could influence progression to D2T RA under real-life conditions. Other clinical and therapeutic factors were also analyzed.

## Methods

### Setting

The study was performed at three tertiary hospitals of the National Health System of the Community of Madrid, Spain, namely, Hospital Clínico San Carlos (HCSC), Hospital Universitario de La Paz (HULP), and Hospital Universitario de la Princesa (HUP). Each institution has a catchment population of approximately 400,000. The Rheumatology Service at each institution provides care to its catchment population.

### Study Design

We performed a multicenter observational longitudinal prospective study. Since we have been treating RA according to the EULAR/ACR classification criteria under a treat-to-target strategy since 2010 (23), patients were included from June 2009 to June 2018 and followed up for five years or less (loss of follow-up or end of the study [January 2021]).

### Population, patients, and data sources

All patients fulfilled the diagnostic criteria for RA and were included at diagnosis. Data were registered prospectively at baseline and every 6-12 months in the local databases and collected in the IntegrAR cohort.

Briefly, the IntegrAR cohort is the product of integrating local prospective inception RA cohorts with standardized data. IntegrAR was initiated in 2010 and contains a relational database whose records are fed from the databases of the three hospitals involved through web services (HCSC, HULP, and HUP). It is an active and dynamic cohort that currently contains more than 210 variables, including sociodemographic and clinical characteristics, diagnostic procedures, and therapeutic options.

To be included, patients had to be in the IntegrAR cohort during the study period, age ≥18 years, and diagnosed with RA according to the EULAR 2010 classification criteria(23). They also had to be taking bDMARDs or tsDMARDs.

Patient data were obtained during routine clinical practice, and patients from IntegrAR provided their signed informed consent to participate. The study was conducted in accordance with the Declaration of Helsinki and Good Clinical Practice and was approved by the local ethics committees (18/321-E_BS)

### Outcome measures

The primary outcome was D2T RA as defined by the EULAR criteria (14), which are set out below.

1. Treatment according to EULAR recommendations and failure of ≥2 bDMARDs/tsDMARDs (with different mechanisms of action) after failure of conventional synthetic DMARDs (unless contraindicated)
2. Presence of at least one of the following: moderate or more severe disease activity (DAS28 ≥3.2), signs and/or symptoms suggestive of active disease (erythrocyte sedimentation rate [ESR] ≥60, C-reactive protein [CRP] ≥0.6), inability to taper glucocorticoids, rapid radiographic progression, and RA symptoms that worsen quality of life (HAQ ≥1.2);
3. The management of signs and/or symptoms is perceived as problematic by the rheumatologist and/or the patient (physician global assessment ≥50; patient global assessment ≥50; text recorded by the rheumatologist in the clinical history). The second and the third criteria had to be fulfilled at least 6 months after diagnosis. The main variable measure in the present study was low disease activity, understood as a DAS28 <3.2.

The covariables initially considered were as follows:

a. Baseline sociodemographic characteristics including center, sex, date of birth, and educational level.
b. Associated baseline comorbidity (hypertension, hypercholesterolemia, diabetes mellitus, cardiovascular and cerebrovascular events, congestive heart failure, depression, history of cancer, gastroduodenal ulcer, chronic obstructive pulmonary disease, interstitial lung disease, liver disease, osteoporosis, kidney failure).
c. Characteristics of RA: date of onset of symptoms, date of diagnosis, date of first visit, rheumatoid factor (RF), anti-citrullinated protein antibody (ACPA).
d. Assessment of RA at baseline: number of painful and swollen joints, assessments of the disease by doctor and patient (patient global assessment, physician/evaluator global assessment), pain scale, DAS28 score, disability (HAQ), ESR.
e. First therapeutic regimen received: DMARDs scheduled with start and end date and glucocorticoids at baseline. The list of DMARDs included csDMARDs (azathioprine, gold, methotrexate, leflunomide, sulfasalazine, and antimalarials) and biologic agents (ts/bDMARDs, TNF inhibitors [anti-TNF], abatacept, rituximab, tocilizumab, and JAK inhibitors). Patients taking any DMARD for more than three months were considered exposed. In the case of glucocorticoids, patients were considered exposed if on medication for at least two months. First, we carried out feature selection, where we combined comorbidity data into two variables: 1) presence of hypertension, hypercholesterolemia, and/or diabetes mellitus; and 2) total number of comorbidities, excluding the previous ones. Second, we selected covariables with a prevalence higher than > 5% in the low-disease-activity group. Finally, only the covariables suspected of playing a relevant role in developing D2T RA were considered to balance the distribution between patients with low disease activity and patients with activity (age, sex, combined therapy, HAQ, RF-positive, ACPA-positive, depression, year of diagnosis, comorbidity, intake of glucocorticoids and methotrexate, and center).

### Statistical Analyses

Descriptive statistics of patients’ sociodemographic and clinical characteristics, as well as their disease activity and treatment, are presented as frequency distributions for qualitative variables and as the mean and standard deviation or median and percentiles for quantitative variables. The *t* test was used for the analysis of normally distributed continuous variables. Continuous variables with a non-normal distribution were analyzed using the Mann-Whitney test or the Kruskal-Wallis test if there were more than two categories. The categorical variables were analyzed using the chi-square or Fisher exact test. Analyses were performed to assess the differences between D2T RA and non–D2T RA. Logistic multivariate regression models were fitted to examine the possible influence of baseline disease activity (low activity) on the main outcome, D2T RA, after balancing the covariates of both groups (i.e., low disease activity and activity groups) using different balancing strategies. These models were adjusted for the independent variables of disease activity (DAS28, HAQ) and therapy (intake of glucocorticoids and methotrexate). We also fitted a logistic multivariate regression model without covariate balancing. Details on the methods used to balance the distribution of variables can be found in the Supplemental file “Statistical analyses”.

We limited the number of variables in the multivariate model following the rule of Freeman and the value of 10 events per variable to ensure that it was reliable(24)(25)(25).

Analyses were performed using STATA v13 software (Stata Corp) and R statistical software version 4.2.1(26). The *WeightIt* (27) and *cobalt* (28) R packages were used to estimate the propensity score weights. Missing data (HAQ n = 27, DAS28 n = 17) were managed using the classification and regression trees algorithm (CART) from the *mice* R package, with the default setting and 5 imputations (29).

## Results

### Sociodemographic, clinical, and therapy-related characteristics

A total of 631 patients were included: 178 (28%) from HCSC, 319 (51%) from HULP, and 134 (21%) from HUP. All of them were taking DMARDs during the study period, and the median follow-up was 5 [p25-p75: 2.14-5] years. Most patients were women (77.5%) in their fifties, with no statistically significant differences between the centers. Lag time from symptoms to diagnosis of RA was 0.7 (2.2) years. Interestingly, 30% of the patients had at least 1 comorbid condition at baseline and 28% had a cardiovascular risk factor, with arterial hypertension being the most prevalent. Most of the patients were overweight, with obesity in 10% (median BMI, 25.8 [p25-p75: 22.7-28.9]). Most were RF- or ACPA-positive, with a median value of 106 [p25-p75 42-285] and 445 [p25-p75: 202-1055], respectively. Regarding disease activity, most patients had at least a moderate DAS28 (median, 4.7 [p25-p75: 3.6-5.8]) and some degree of disability (median HAQ, 1 [0.375-1.625]).

Glucocorticoids were the first drugs prescribed in 66% of the patients. Most patients were prescribed csDMARDs at diagnosis (71.54%), 97% in the first six months, and 99% in the first 12 months, with a mean start of 30.41 (65) days after diagnosis. The drugs were prescribed in monotherapy in 87% of the cases, and, as expected, the most prevalent was methotrexate (85%) either in combination or in monotherapy. Ten patients (1.58%) received ts/bDMARDs as their initial treatment, with etanercept (n=3) and abatacept (n=3) the most frequently prescribed, followed by tocilizumab (n=2), adalimumab (n=1), and golimumab (n=1). The remaining patients were prescribed methotrexate, leflunomide, or hydroxychloroquine, except for one patient who received etanercept.

Interestingly, 17% of patients used at least one ts/bDMARDs during follow-up, with a median first prescription lag time of 1.4 [0.79-2.5] years after diagnosis. Of these, 42 patients required at least two different biologics during the study period. Concerning the other D2T RA criteria, 63% and 39% of patients met the second and third criteria, respectively (see Figure 1). Finally, 35 cases were defined as D2T RA and 596 cases as non–D2T RA. Clinical characteristics and therapy are summarized in Table 1 and Table 2.

**Table 1.** Baseline characteristics of patients in the D2T RA and the non–D2T RA groups

| Variable | RA<br>n=631 | D2T RA<br>n=35 | Non-D2T RA<br>n=596 | p |
| --- | --- | --- | --- | --- |
| Women, n (%) | 489 (77.50) | 29 (82.86) | 460 (77.18) | 0.4 |
| Age, mean (SD), years | 55.10 (15.57) | 45.79 (12.89) | 55.64 (15.55) | 0.0003 |
| BMI, mean (SD) | 26.09 (4.72) | 26.96 (4.09) | 26.04 (4.75) | 0.37 |
| Positive RF at baseline, n (%) | 526 (83.36) | 24 (68.57) | 502 (84.23) | 0.01 |
| Positive ACPA at baseline, n (%) | 499 (79.08) | 26 (74.29) | 473 (79.36) | 0.47 |
| DAS28 at RA diagnosis, mean (SD) | 4.68 (1.51) | 5.08 (1.43) | 4.66 (1.52) | 0.11 |
| CRP at RA diagnosis, mean (SD) | 1.98 (5.91) | 1.51 (2.10) | 2.01 (6.06) | 0.65 |
| ESR at RA diagnosis, mean (SD) | 30.05 (22.29) | 28.36 (22.25) | 30.15 (22.31) | 0.65 |
| TJC (0-28) at baseline, mean (SD) | 6.73 (6.55) | 8.94 (7.98) | 6.60 (6.44) | 0.04 |
| SJC (0-28) at baseline, mean (SD) | 6.60 (5.81) | 7.71 (6.55) | 6.53 (5.76) | 0.24 |
| PGA (mm) at baseline, mean (SD) | 46.93 (26.53) | 59.2 (26.90) | 46.19 (26.34) | 0.004 |
| PhGA (mm) at baseline, mean (SD) | 38.71 (22.18) | 43.85 (20.21) | 38.47 (22.25) | 0.29 |
| Pain VAS (mm) at baseline, mean (SD) | 45.24 (28.04) | 56.59 (27.73) | 44.16 (1.65) | 0.02 |
| HAQ at RA diagnosis, mean (SD) | 1.04 (0.74) | 1.30 (0.66) | 1.02 (0.74) | 0.03 |
| Comorbidities, n (%) |  |  |  |  |
| Hypertension | 124 (19.65) | 5 (14.29) | 119 (19.97) | 0.41 |
| Dyslipidemia | 93(14.74) | 10 (28.57) | 83 (13.93) | 0.018 |
| Depression | 36(5.71) | 4 (11.43) | 32 (5.37) | 0.13 |
| Diabetes mellitus | 34(5.39) | 2 (5.72) | 32 (5.37) | 0.96 |
| Heart disease | 14 (2.22) | 0 | 14 (2.35) | 0.35 |
| Vascular disease | 33 (5.22) | 1 (0.15) | 32 (5.07) | 0.72 |
| Liver disease | 8 (1.27) | 2 (5.72) | 6 (1.01) | 0.0001 |
| Kidney disease | 4 (0.63) | 0 | 4 (0.67) | 0.62 |
| Lung disease (ILD/COPD) | 18 (2.85) | 1 (2.86) | 17 (2.85) | 0.99 |
| Cancer | 8 (4.49) | 0 | 8 (4.97) | 0.34 |

**Abbreviations:** D2T RA: difficult-to-treat rheumatoid arthritis; SD: standard deviation; BMI: body mass index; RF: rheumatoid factor; ACPA: anti-citrullinated-protein antibody; DAS28: 28-joint disease activity score; CRP: C-reactive protein; ESR: erythrocyte sedimentation rate; TJC: tender joint count; SJC: swollen joint count; PGA: patient global assessment, PhGA: physician global assessment; VAS: visual analog score; HAQ: Health Assessment Questionnaire; ILD: interstitial lung disease; COPD: chronic obstructive pulmonary disease.

**Table 2.**
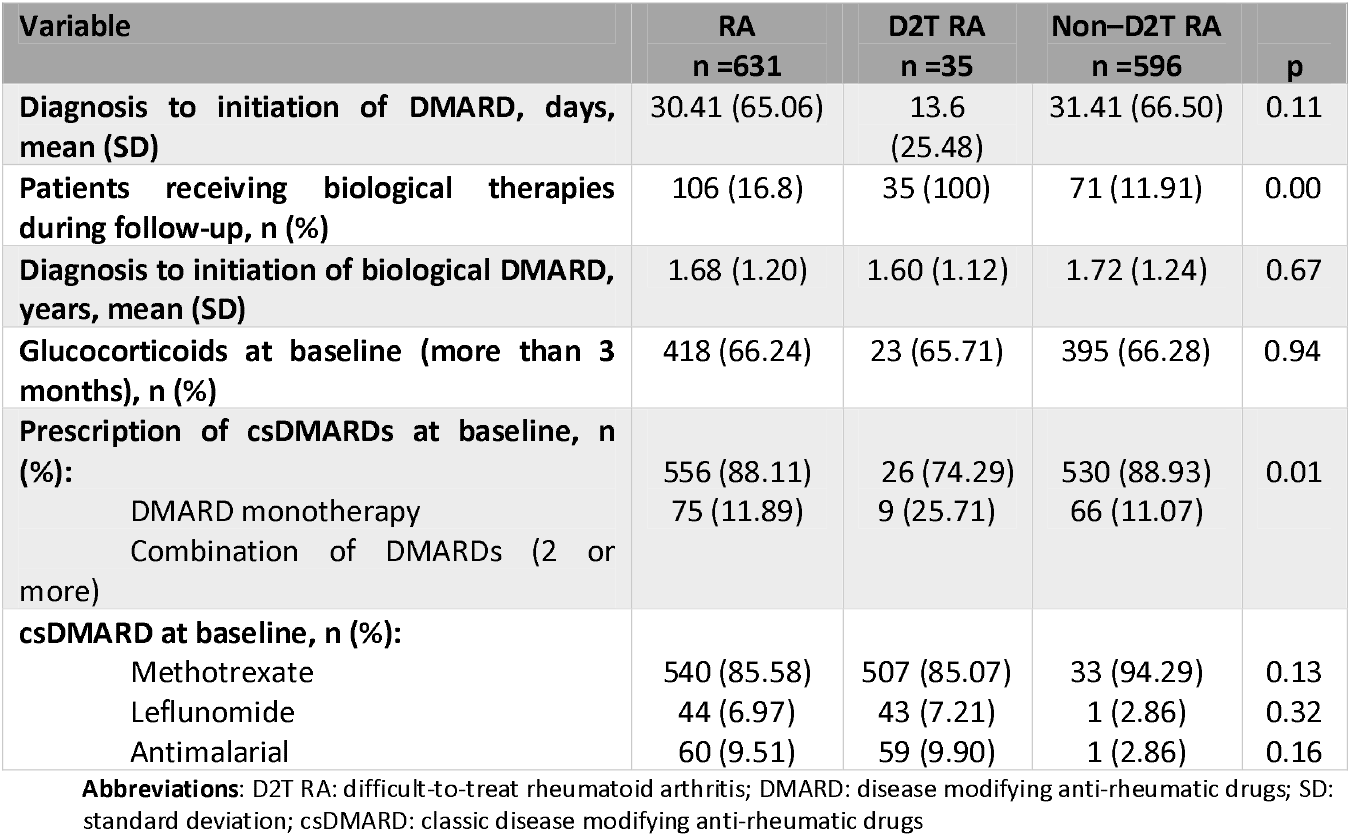
Baseline treatment strategies in the D2T RA and the non–D2T RA groups

**Figure 1.**
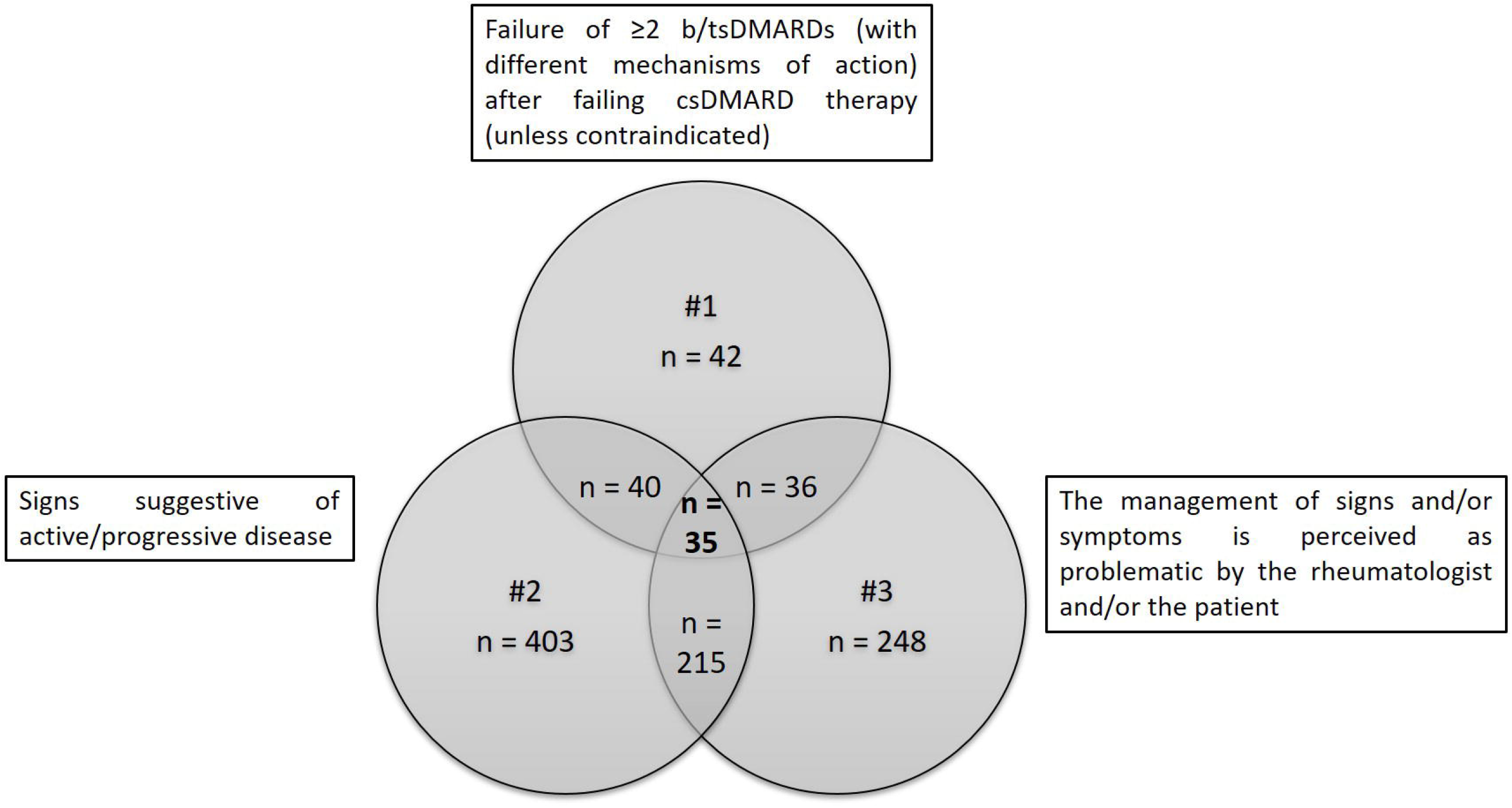
Number of patients who met each of the three D2T RA criteria.

At diagnosis, patients in the D2T RA group were younger, with a higher DAS28 tender joint count, higher pain scores, and a higher level of disability than the non–D2T RA group. Regarding comorbidities, the main differences were observed for dyslipidemia (28% vs 13%) and liver disease (5% vs 1%), with a greater prevalence in D2T RA patients, in whom a trend toward depression was observed (11% vs 5%). Other characteristics are shown in Table 1.

Table 2 shows differences in therapy between the 2 groups. No statistically significant differences were recorded between the groups for prevalence of the different DMARDs or glucocorticoids. As expected, D2T RA patients took more bDMARDs during follow-up. Regarding the biologics used as the initial regimen, all except one taking abatacept were in the non–D2T RA group.

### Influence of baseline disease activity on D2T RA

Supplemental Tables S1 and S2 show the mean differences for the selected covariables— unadjusted and weighted (adjusted, trimmed, and stabilized)—using methods based and not based on the propensity score (PS), respectively.

Only methods not based on the propensity score, entropy balancing (EBAL), and empirical balancing calibration weighting (EBCW) were able to balance all the covariates. We selected EBAL with stabilized weights, as this configuration was associated with the lowest coefficient of variation.

In the weighted multivariable logistic regression analysis (Table 3), we did not find statistically significant differences for disease activity (OR 1.8 [0.5-6.7], p=0.46), methotrexate (OR 2.3 [0.5-10.2], p=0.27), or glucocorticoids (OR 1.02[0.5-2.2], p=0.98), whereas at least moderate disability (OR 1.5 [1.1-2.1], p=0.01) increased the risk of D2T RA, regardless of other factors.

**Table 3:** Multivariate regression analysis

|  | OR | 95% CI | p |
| --- | --- | --- | --- |
| **DAS28** | 1.82 | 0.5-6.67 | 0.36 |
| **Glucocorticoids** | 1.02 | 0.46-2.23 | 0.96 |
| **Methotrexate** | 2.35 | 0.54-10.2 | 0.25 |
| **Health Assessment Questionnaire** | 1.50 | 1.07-2.09 | 0.02 |

**Abbreviations:** DAS28: 28-joint disease activity score; OR: odds ratio
Analysis balanced for age, sex, center, disability, rheumatoid factor, anti-citrullinated-protein antibody, depression, year of diagnosis, comorbidity, disease modifying anti-rheumatic drugs, and glucocorticoids.

Similar results were found in the unweighted multivariable logistic regression analysis, as follows: disease activity (OR 1.1 [0.4-3], p=0.85), methotrexate (OR 2.8 [0.8-17.6], p=0.16), glucocorticoids (OR 0.9[0.4-1.9], p=0.79), and at least moderate disability (OR 1.4 [0.9-2], p=0.04).

## Discussion

In this study, we investigated whether early disease activity plays a role in the detection of D2T RA under real-world conditions. Our results do not allow us to prove the influence of active disease state according to DAS28, although we did find that patients with elevated initial disability scores are more likely to develop D2T RA, regardless of other factors. Our evaluation of whether the first therapeutic strategy might result in better outcomes depending on the drugs selected did not yield conclusive results.

As in other studies, recent-onset RA patients were middle-aged women whose most common clinical and serological manifestations were positive RF and ACPA titers (30)(31).

We found that 5.87% of patients had D2T RA, consistent with findings reported elsewhere (15)(20). Previous studies of D2T RA patients were based only on treatment failure, without taking into account other criteria (i.e. signs suggestive of presently active/progressive disease and management being perceived as problematic by the rheumatologist and/or patient). Although several studies have described D2T RA–related factors in RA patients (18)(32)(33) with long-standing disease, the effect of different variables on this outcome has not been studied in the early stages of disease, and comparisons between them would not be accurate.

Although an evidence base is not available, identifying affected patients in the early stages of disease could, arguably, limit progression to D2T RA in those experiencing multi-drug failure. In our study, we systematically analyzed the effect of several variables, mainly those related to disease; however, we also examined the response to treatment (DMARDs and glucocorticoids).

Disease activity (moderate, severe, early) based on the DAS 28 was not associated with D2T RA, although statistically significant differences were observed after separate analysis of DAS28 components, tender joint score, and patient global assessment, suggesting that rheumatologists act well towards the most objective part of the disease, with little emphasis on the more subjective aspects (34). Interestingly and similarly, a statistically significant higher early disease burden was found in D2T RA than in non D2T RA patients, as seen in lower physical functioning and worse levels of pain; therefore, it was not possible to distinguish disease activity arising from pain or inflammation per se. While temporary, this finding has been reported in other studies of patients with much longer-standing disease who eventually develop D2T RA (18). Nevertheless, structural and irreversible disability may develop until inflammation is controlled, with the result that early control of function should remain a mandatory objective.

As for the “dual-target” strategy suggested in response to the risks of overtreatment with immunosuppression in pursuit of a treatment target that may not be achievable with drug therapy alone, Ferreira et al (35) recommend a personalized approach that also addresses the impact of the disease on the patient, including disability, which can be assessed using patient-reported outcome measures and requires a multidisciplinary approach.

Several non-pharmacological therapeutic strategies have proven helpful in RA patients. Educational, psychological, and self-management interventions have diminished non-inflammatory complaints (pain, fatigue, and functional disability)(36)(37)(38), and while high-quality evidence of the beneficial effect of these non-pharmacological strategies is still lacking in D2T RA patients, they could prevent a large number of cases of D2T RA.

A novel aspect of the present study was our evaluation of the role of different treatment strategies in the development of D2T RA. We were unable to find differences between RA groups with respect to therapeutic management. The use of glucocorticoids does not seem to have any effect, as reported in a sample of patients with long-standing D2T RA. (22). Similarly, the choice of methotrexate as the first DMARD does not seem to affect the future development of D2T RA. In a recent study, D2T RA patients receiving methotrexate⍰≤8.6 mg at baseline are considered to be at high risk of recurrence of D2T RA (22). The authors reported that their doses were lower than in Western countries. The mean dose we prescribe after three months is around 15-20 mg/week (common practice is to start with a lower dose, (39)); however, data were not collected systematically, thus precluding comparisons.

Methotrexate is the cornerstone of treatment of early RA, although it is likely that higher doses are more poorly tolerated. Despite common use of folic acid, intolerance to methotrexate is still reported in 30%-60% of patients (40)(41). This may be one of the reasons why adherence is only 50%-94% at 1 year and 25%-79% at 5 years (42). Intolerance to methotrexate was associated with younger age, as reported elsewhere (41)(43)(44). In our study, fewer patients with D2T RA took methotrexate than in the non–D2T RA group. The age difference between the groups may have affected the use and retention of methotrexate.

Early introduction of aggressive immunosuppressive treatment at appropriate dosages and with combination therapy when needed is recommended for achieving remission (45). We found no differences between the lag time to the first DMARDs used, although most were prescribed early, suggesting that the “treat to target” strategy and the current EULAR recommendations are being implemented in clinical practice, thus leading to good control of the disease. Consequently, given that disease control has improved, disease activity is probably not an associated factor.

Previous studies questioned whether the sequence of DMARDs and/or resistance to specific drugs affects progression to D2T RA (21). Our analysis of drug choice and combinations showed no trend toward D2T RA. One reason why the first regimen is not a determinant in this condition could be that rheumatologists are applying the treat-to-target strategy in clinical practice and that modifications (adding, switching) are made so quickly that the effect of the initial regimen is diminished. Another reason might be that the most frequently prescribed drug is methotrexate, making it difficult to find differences between treatments. Lastly, we cannot forget that the D2T RA group is too small and heterogeneous in terms of therapy to found differences.

Our study is subject to a series of limitations. First, its observational nature meant that treatment was not allocated randomly, and although we used various methods to increase the comparability of both groups, prescription bias cannot be completely ruled out. However, we observed similar results between the balancing method chosen and the non-balanced multivariable logistic regression analysis. Consequently, we are confident about the validity of our results. Second, as data were collected from clinical practice, some were missing; however, these account for no more than 5%, and imputation methods were applied to correct this issue. Third, given the extended follow-up, some of the data may reflect clinical practice that has changed over time as more therapeutic options have become available. We included cases treated several years ago, when biologic agents were not available. Once they did become available, combinations were not usual until experience and evidence started to support them. This is also reflected in our limited experience with other targeted therapies, such as JAK inhibitors. The balanced methods including year of diagnosis of RA enabled us to correct this bias. Finally, despite the 10-year inclusion period, the number of events was small, thus diminishing the strength of the analysis in terms of, for example, therapeutic regimen used. Nevertheless, the strengths of our study include its prospective design and the inclusion of non-selected patients from three hospitals with a follow-up limited to five years, which provided us with an overall vision of real-world evidence and thus reflected clinical practice in our setting. Our results add to current knowledge on the real-world management of D2T RA in the window of opportunity. In addition, more than 600 patients were analyzed, and several weighting techniques applied, with consistent results.

Our findings should be interpreted as an initial approach to the potential role that early detection of clinical issues could play in the prevention of D2T RA. A shift in current clinical practice is needed to encourage clinicians to consider comorbidities and predisposing conditions early in the disease, before D2T RA develops, rather than after it manifest (46). Other observational studies, starting at disease onset, will be needed to resolve these issues.

To conclude, the implementation of more effective strategies in the early stages of the disease and focused on the most influential factors, including severe disability, may change disease course and prevent D2T RA. Nevertheless, the presence of refractory disease highlights the need for continued target discovery, new therapies, and a multidisciplinary approach.

## Data Availability

The datasets used and/or analyzed during the current study are available from the corresponding author on reasonable request

