## Supplemental Table 1 for "Difficult-to-treat rheumatoid arthritis (D2T RA): clinical issues at early stages of disease"

**Table S1**: Mean differences for the covariates included in the analysis of the effect of low baseline disease activity in D2T RA before and after application of propensity score–based methods.

|  |  | GLM | | | GBM | | | CBPS | | | NPCBPS | | | MLPS | | |
| --- | --- | --- | --- | --- | --- | --- | --- | --- | --- | --- | --- | --- | --- | --- | --- | --- |
| Covariates | Unadj | Adj | Trim | Stab | Adj | Trim | Stab | Adj | Trim | Stab | Adj | Trim | Stab | Adj | Trim | Stab |
| Age | -0.112 | 0.004 | 0.005 | 0.004 | -0.034 | -0.036 | -0.034 | 0.005 | 0.001 | 0.005 | -0.028 | -0.013 | -0.028 | -0.075 | -0.073 | -0.075 |
| Women | -0.085 | -0.008 | -0.025 | -0.008 | -0.025 | -0.039 | -0.025 | -0.011 | -0.024 | -0.011 | -0.025 | -0.03 | -0.025 | -0.022 | -0.033 | -0.022 |
| Combined therapy | -0.079 | -0.035 | -0.03 | -0.035 | -0.077 | -0.073 | -0.077 | -0.018 | -0.019 | -0.018 | -0.02 | -0.021 | -0.02 | -0.077 | -0.074 | -0.077 |
| HAQ | -1.179 | -0.246 | -0.416 | -0.246 | -0.51 | -0.611 | -0.51 | -0.367 | -0.477 | -0.367 | -0.316 | -0.39 | -0.316 | -0.666 | -0.746 | -0.666 |
| RF | 0.029 | -0.007 | 0.022 | -0.007 | 0.026 | 0.017 | 0.026 | 0.017 | 0.029 | 0.017 | 0.01 | -0.001 | 0.01 | 0.029 | 0.021 | 0.029 |
| ACPA | 0.092 | -0.002 | 0.031 | -0.002 | 0.088 | 0.083 | 0.088 | 0.02 | 0.033 | 0.02 | 0.026 | 0.02 | 0.026 | 0.082 | 0.074 | 0.082 |
| Depression | 0.017 | 0.044 | 0.007 | 0.044 | 0.012 | 0.017 | 0.012 | 0.02 | 0.003 | 0.02 | 0.004 | 0.01 | 0.004 | 0.013 | 0.017 | 0.013 |
| Year | 0.048 | -0.012 | -0.041 | -0.012 | -0.043 | -0.092 | -0.043 | 0.012 | -0.014 | 0.012 | 0.018 | -0.021 | 0.018 | -0.017 | -0.01 | -0.017 |
| Number of comorbidities | -0.102 | -0.167 | -0.119 | -0.167 | -0.135 | -0.107 | -0.135 | -0.144 | -0.109 | -0.144 | -0.032 | 0 | -0.032 | -0.125 | -0.129 | -0.125 |
| Dyslipidemia | 0.022 | 0.047 | 0.019 | 0.047 | 0.017 | 0.026 | 0.017 | 0.027 | 0.009 | 0.027 | 0.007 | 0.023 | 0.007 | 0.013 | 0.013 | 0.013 |
| Corticosteroid | -0.165 | 0.041 | 0.011 | 0.041 | -0.058 | -0.067 | -0.058 | 0.014 | -0.008 | 0.014 | -0.042 | -0.058 | -0.042 | -0.047 | -0.063 | -0.047 |
| Center HULP | -0.007 | 0.014 | 0.021 | 0.014 | 0.063 | 0.035 | 0.063 | 0.019 | 0.019 | 0.019 | -0.001 | -0.016 | -0.001 | 0.045 | 0.021 | 0.045 |
| Center HCSC | -0.028 | -0.055 | -0.034 | -0.055 | -0.075 | -0.059 | -0.075 | -0.03 | -0.019 | -0.03 | -0.006 | -0.001 | -0.006 | -0.051 | -0.041 | -0.051 |
| Center HUP | 0.035 | 0.041 | 0.013 | 0.041 | 0.012 | 0.024 | 0.012 | 0.012 | 0 | 0.012 | 0.008 | 0.017 | 0.008 | 0.006 | 0.019 | 0.006 |
| Methotrexate | -0.02 | 0.042 | 0.031 | 0.042 | 0.025 | 0.016 | 0.025 | 0.034 | 0.026 | 0.034 | 0.028 | 0.021 | 0.028 | 0.018 | 0.01 | 0.018 |
| Number of  unweighted covariates | - | 3 | 2 | 3 | 7 | 7 | 7 | 2 | 2 | 2 | 1 | 2 | 1 | 6 | 6 | 6 |
| Coefficient of  variance | - | 1.539 | 1.183 | 0.509 | 0.979 | 0.790 | 0.349 | 1.318 | 1.105 | 0.430 | 0.574 | 0.472 | 0.414 | 0.680 | 0.548 | 0.295 |

**Abbreviations**:Adj: adjusted; CBPS: covariate balancing method; GBM: generalized boosted models; GLM: generalized linear models; HULP: Hospital Universitario de La Paz; HCSC: Hospital Clínico San Carlos; HUP: Hospital Universitario de la Princesa; MLPS: machine learning–based PS methods; NPCBPS: non-parametric covariate balancing method; Stab: stabilized; Trim: trimmed; Unadj: unadjusted RF: rheumatoid factor; ACPA: anti-citrullinated protein antibody.
