## Supplemental Table 2 for "Difficult-to-treat rheumatoid arthritis (D2T RA): clinical issues at early stages of disease"

**Table S2**: Mean differences for the covariates included in the analysis of the effect of low baseline disease activity in D2T RA before and after application of non-propensity score–based methods.

|  | EBAL | | | | | EBCW | | | | OBW | | | | Energy | | |
| --- | --- | --- | --- | --- | --- | --- | --- | --- | --- | --- | --- | --- | --- | --- | --- | --- |
| Covariates | Unadj | Adj | Trim | Stab | Adj | | Trim | Stab | Adj | | Trim | Stab | Adj | | Trim | Stab |
| Age | -0.112 | -2.69E-05 | 0.025 | -2.69E-05 | 2.61E-10 | | 0.025 | 2.61E-10 | 8.45E-14 | | 0.001 | 8.45E-14 | -0.024 | | -0.025 | -0.024 |
| Women | -0.085 | 3.08E-05 | -0.011 | 3.08E-05 | -2.58E-10 | | -0.011 | -2.58E-10 | 4.73E-14 | | -0.001 | 4.72E-14 | -0.001 | | 0 | -0.001 |
| Combined therapy | -0.079 | -3.95E-05 | 0.002 | -3.95E-05 | -1.16E-10 | | 0.002 | -1.16E-10 | 5.04E-14 | | -0.02 | 5.04E-14 | -0.01 | | -0.009 | -0.01 |
| HAQ | -1.179 | 2.63E-06 | -0.053 | 2.63E-06 | -1.87E-09 | | -0.053 | -1.87E-09 | -1.123 | | -1.116 | -1.123 | -0.174 | | -0.194 | -0.174 |
| RF | 0.029 | 6.82E-05 | -0.006 | 6.82E-05 | 7.57E-11 | | -0.007 | 7.57E-11 | -1.48E-14 | | -0.003 | -1.49E-14 | 0.002 | | 0.002 | 0.002 |
| ACPA | 0.092 | 8.91E-06 | -0.008 | 8.91E-06 | 1.91E-10 | | -0.008 | 1.91E-10 | -5.98E-14 | | 0.011 | -6.00E-14 | 0.003 | | 0.003 | 0.003 |
| Depression | 0.017 | -7.81E-06 | 0.001 | -7.81E-06 | 2.17E-11 | | 0.001 | 2.17E-11 | -1.15E-14 | | 0.001 | -1.15E-14 | 0.003 | | 0.003 | 0.003 |
| Year | 0.048 | 1.94E-05 | -0.046 | 1.94E-05 | 2.06E-11 | | -0.046 | 2.06E-11 | 0.06 | | 0.052 | 0.06 | 0.005 | | -0.028 | 0.005 |
| Number of comorbidities | -0.102 | -3.95E-05 | 0.026 | -3.95E-05 | -1.30E-10 | | 0.026 | -1.30E-10 | -0.102 | | -0.105 | -0.102 | -0.03 | | -0.014 | -0.03 |
| Dyslipidemia | 0.022 | -5.48E-06 | 0.013 | -5.48E-06 | 1.73E-10 | | 0.013 | 1.73E-10 | -5.27E-15 | | 0.001 | -5.33E-15 | 0.001 | | 0.001 | 0.001 |
| Corticosteroid | -0.165 | -5.74E-05 | -0.005 | -5.74E-05 | -4.72E-10 | | -0.005 | -4.72E-10 | 8.74E-14 | | -0.003 | 8.75E-14 | -0.001 | | -0.007 | -0.001 |
| Center HULP | -0.007 | -2.25E-05 | -0.02 | -2.25E-05 | -4.48E-11 | | -0.02 | -4.48E-11 | -9.44E-15 | | 0.01 | -9.44E-15 | -0.001 | | -0.008 | -0.001 |
| Center HCSC | -0.028 | -1.69E-06 | 0.011 | -1.69E-06 | 9.62E-12 | | 0.011 | 9.62E-12 | 1.64E-14 | | -0.013 | 1.63E-14 | 0 | | 0 | 0 |
| Center HUP | 0.035 | 2.42E-05 | 0.009 | 2.42E-05 | 3.52E-11 | | 0.009 | 3.52E-11 | -6.94E-15 | | 0.003 | -6.97E-15 | 0 | | 0.007 | 0 |
| Methotrexate | -0.02 | 0.048 | 0.043 | 0.048 | 0.048 | | 0.043 | 0.048 | -0.008 | | 0.004 | 0.008 | -0.014 | | -0.014 | -0.014 |
| Number of  unweighted  covariates | - | 0 | 1 | 0 | 0 | | 1 | 0 | 3 | | 3 | 3 | 1 | | 1 | 1 |
| Coefficient of  variance | - | 0.731 | 0.662 | 0.446 | 2.168 | | 1.996 | 0.731 | 0.238 | | 0.213 | 0.372 | 1.065 | | 0.983 | 1.156 |

**Abbreviations** Adj: adjusted; ACPA: anti-citrullinated protein antibody; EBAL: entropy balancing; EBCW: empirical balancing calibration weighting; HAQ: Health Assessment Questionnaire; HULP: Hospital Universitario de La Paz; HCSC: Hospital Clínico San Carlos; HUP: Hospital Universitario de la Princesa; RF: rheumatoid factor; OBW: optimization-based weighting; Stab: stabilized; Trim: trimmed; Unadj: unadjusted.
