## Supplemental Table 3 for "Difficult-to-treat rheumatoid arthritis (D2T RA): clinical issues at early stages of disease"

**Table S3**: Multivariable logistic regression results of the methods that achieve full covariate balance

|  | Adjusted | | | Stabilized | | |
| --- | --- | --- | --- | --- | --- | --- |
| **EBAL** | **OR** | **95% CI** | **p-value** | **OR** | **95% CI** | **p-value** |
| DAS28 | 1.866 | 0.004-0.096 | 0.354 | 1.82 | 0.501-6.673 | 0.36 |
| HAQ | 1.752 | 0.498-6.997 | 0.030 | 1.50 | 1.074-2.096 | 0.017 |
| Glucocorticoid | 1 | 0.427-2.3489 | 0.997 | 1.02 | 0.467-2.232 | 0.958 |
| Methotrexate | 2.646 | 0.609-11.478 | 0.194 | 2.35 | 0.545-10.174 | 0.25 |
| **EBCW** | **OR** | **95% CI** | **p-value** | **OR** | **95% CI** | **p-value** |
| DAS28 | 2.082 | 0.002-0.09 | 0.338 | 1.867 | 0.498-7 | 0.354 |
| HAQ | 2.675 | 0.464-9.346 | 0.104 | 1.752 | 1.056-2.907 | 0.03 |
| Glucocorticoid | 0.888 | 0.204-3.872 | 0.874 | 1.002 | 0.427-2.349 | 0.997 |
| Methotrexate | 3.554 | 0.777-16.250 | 0.102 | 2.654 | 0.609-11.478 | 0.194 |

**Abbreviations**: EBAL: entropy balancing; DAS28, 28-joint Disease Activity Score; HAQ: Health Assessment Questionnaire; EBCW: empirical balancing calibration weighting; OR: odds ratio; CI: confidence interval
